# SARS-CoV-2 genomic surveillance enables the identification of Delta/Omicron co-infections in Argentina

**DOI:** 10.1101/2022.03.08.22270920

**Authors:** María Belén Pisano, Paola Sicilia, Maximiliano Zeballos, Andrea Lucca, Franco Fernandez, Gonzalo M. Castro, Stephanie Goya, Mariana Viegas, Laura López, María Gabriela Barbás, Viviana E. Ré

**Affiliations:** Instituto de Virología “Dr. J. M. Vanella”, CONICET, Facultad de Ciencias Médicas, Universidad Nacional de Córdoba. Enfermera Gordillo Gómez s/n, Ciudad Universitaria, CP5016 Córdoba, Argentina; Consejo Nacional de Investigaciones Científicas y Técnicas (CONICET), Argentina; Laboratorio Central de la Provincia de Córdoba, Ministerio de Salud de la Provincia de Córdoba. Tránsito Cáceres de Allende 421, Córdoba, Argentina; Fundación para el Progreso de la Medicina; Instituto Nacional de Tecnología Agropecuaria (INTA), Centro de Investigaciones Agropecuarias (CIAP), Instituto de Patología Vegetal (IPAVE). Camino 60 cuadras km 5 ½ (X5020ICA), Córdoba, Argentina; Consejo Nacional de Investigaciones Científicas y Técnicas (CONICET). Unidad de Fitopatología y Modelización Agrícola (UFYMA). Camino 60 cuadras km 5 ½ (X5020ICA), Córdoba. Argentina; Laboratorio de Virología, Hospital de Niños Dr. Ricardo Gutiérrez, Buenos Aires, Argentina; Área de Epidemiología, Ministerio de Salud de la Provincia de Córdoba. Av. Vélez Sarsfield 2311 Ciudad Universitaria, X5016 GCH, Córdoba, Argentina; Secretaría de prevención y promoción de la salud, Ministerio de Salud de la Provincia de Córdoba. Av. Vélez Sarsfield 2311 Ciudad Universitaria, X5016 GCH, Córdoba, Argentina

**Keywords:** SARS-CoV-2, co-infection, Delta, Omicron, Argentina

## Abstract

Molecular surveillance of SARS-CoV-2 is crucial to early detect new variants and lineages. In addition, detection of coinfections with more than one SARS-CoV-2 lineage have been sporadically reported. In this work, surveillance of SARS-CoV-2 variants was performed on 2067 RNA samples (Ct>30) obtained during December 2021 and January 2022 from Córdoba province, Argentina, by real time RT-PCR specific for VOC/VOI relevant mutations (TaqMan™ SARS-CoV-2 Mutation Panel, Applied Biosystems). The following distribution of variants was obtained: Omicron (54.9%), Delta (44.2%) and Lambda (0.8%). Three samples (0.1%), obtained the last week of December, presented a profile compatible with a Delta/Omicron co-infection. One of them was sequenced by NGS-Illumina, obtaining reads for both VOCs. One of the studied patients presented severe symptoms, although he was not vaccinated and presented risk factors (older than 60 years, arterial hypertension). We describe for the first time in Argentina, the identification of cases of co-infection with two SARS-CoV-2 lineages, VOCs Delta and Omicron, during the third COVID-19 wave in the country (a high viral circulation period), when Delta and Omicron co-circulated. Our findings highlight the importance of continuing with molecular surveillance and co-detection studies of VOC/VOIs, in order to elucidate possible recombination events and the emergence of new variants.

## Introduction

During the two years of the COVID-19 pandemic, the original SARS-CoV-2 that was identified at the end of 2019 has evolved into various lineages (He et al. 2021), presenting characteristic mutations. Among them, variants that posed an increased risk to global public health have been identified as variants of interest (VOI) and variants of concern (VOC), which present a defined pattern of mutations (WHO 2022). Five VOCs -Alpha (lineage B.1.1.7), Beta (lineage B.1.351), Gamma (lineage P.1), Delta (lineage B.1.617.2) and Omicron (lineage B.1.1.529)-, and two VOIs -Lambda (C.37) and Mu (B.1.621)-have been reported up to date (WHO 2022).

Whole genome sequencing (WGS) has been widely used since the beginning of the pandemic to monitor virus variants, to get a better understanding of the virus biology and epidemiology (Hosch et al. 2022). However, it is a time-consuming and expensive technique, that requires trained staff and specific equipment, restricting its access in resource-limited settings (Blairon et al. 2021). As an alternative, reverse transcription real time polymerase chain reactions (RT real time PCR) assays for detection of relevant mutations associated with SARS-CoV-2 variants have been developed, to typify circulating variants, being a more accessible tool for the monitoring of VOCs (Ong et al. 2021, Blairon et al. 2021).

Molecular SARS-CoV-2 surveillance has allowed to identify the simultaneous infection (co-infection) of a single individual by two distinct SARS-CoV-2 lineages, an event that has been sporadically reported (Dezordi et al. 2021, Zhou et al. 2021, Hosch et al. 2022). These cases constitute an opportunity for viral genetic recombination and the emergence of new lineages with differential phenotype (Dezordi et al. 2021), which may cause more severe clinic symptoms (Zhou et al. 2021). The frequency of co-infected patients and its role to promote recombination-driven SARS-CoV-2 evolution is still unknown and poorly understood (Dezordi et al. 2021).

In Argentina, the profile of circulating lineages and variants has been changing throughout the pandemic, as has happened in the rest of the world (Outbreak info 2022). Molecular surveillance in the country started with WGS carried out by the Ministries of Science and Technology, and Health, at the national level (PAIS 2022, Ministerio de Salud de la Nacion 2022). But then, given the increase in the number of COVID-19 cases, the appearance of VOC/VOIs and the need for rapid results that enable the public health decision-making, some provinces implemented different strategies based on real-time RT-PCRs for detection of VOC/VOIs relevant mutations, as additional techniques to the WGS. This was the case of the province of Córdoba, in the central region of the country, where a strategy that combined detection of point mutations was developed (Castro et al. 2021). This strategy served for molecular surveillance of SARS-CoV-2 variants during 2021, enabling the typing of a greater number of samples in less processing time (Castro et al. 2021, Gobierno de la provincia de Cordoba 2022). Moreover, this strategy allowed the detection of Omicron variant for the first time in Cordoba on December 2021, when the third wave of COVID-19 started in the country, displacing VOC Delta, the major variant circulating at that time (Gobierno de la provincia de Córdoba 2022).

In this report, we describe for the first time in Argentina, the identification of cases of co-infection with two SARS-CoV-2 lineages, particularly with VOCs Delta and Omicron, occurred in December 2021, detected within the molecular surveillance carried out during the third COVID-19 wave in the country.

## Materials and methods

### Samples obtained during SARS-CoV-2 genomic surveillance

A total of 2067 SARS-CoV-2 RNA positive samples obtained from oropharyngeal swabs collected during December 2021 and January 2022 in the province of Córdoba (central area of Argentina) were analyzed for VOC/VOI detection as part of molecular surveillance program of the local Government of the Province. The samples had originally been extracted with MegaBio plus Virus RNA Purification Kit II (BioFlux) on the GenePure Pro Nucleic Acid Purification System NPA-32P and amplified by real time RT-PCR using DisCoVery SARS-CoV-2 Nucleic Acid Detection Kit.

### Detection of VOCs by real time RT-PCR

A strategy of SARS-CoV-2 VOC/VOI typing was implemented in the province of Cordoba, Argentina, during 2021, which varied throughout the year according to the the local profile of circulating variants (Castro et al. 2021). By the beginning of December 2021, with a major presence of Delta circulating in the community, the strategy included a first screening of L452R and P681R mutations, followed by detections of P681H, K417N, L452Q (the second round of mutation detection was carried out depending on the results of the first screening) (Castro et al. 2021, Gobierno de la provincia de Cordoba 2022). TaqMan™ SARS-CoV-2 Mutation Panel (Applied Biosystems) was used for detection of the relevant mutations/deletions. Each reaction of real time RT-PCR is performed as multiplex, including probes detecting the wildtype (wt) as well as probes detecting the mutant nucleotide sequences, used to enable detection of wildtype and mutated sequence simultaneously. Briefly, 7µL of RNA were added to 8 µL of a mixture containing TaqPath™ 1-Step RT-qPCR Master Mix, CG (4X), TaqMan™ SARS-CoV-2 Mutation Panel Assay (40X) and nuclease-free water.

### Whole genome sequencing

Samples that resulted compatible with a co-infection profile in the real-time RT-PCRs for VOC/VOI screening, were subjected to WGS by Illumina platform, using the Illumina COVIDSeq RUO kit, version COVIDSeq Test Kit. Manual inspection of variant-specific mutation sites were accessed using the program Tablet (Milne et al. 2013). Sequenced sample was submitted to GISAID database under the accession number EPI_ISL_8938300.

## Results

From the 2067 samples analyzed using the mutation-specific real-time PCR strategy for detection of VOCs/VOIs, 913 (44.2%) belonged to VOC Delta, 1135 (54.9%) to VOC Omicron, 16 (0.8%) to VOI Lambda and 3 (0.1%) presented profiles compatible with co-infections. The distribution of variants varied throughout the 2 months studied, with an abrupt change in viral circulation, being Omicron all detections performed at the end of January 2022 (Figure 1).

**Figure 1.**
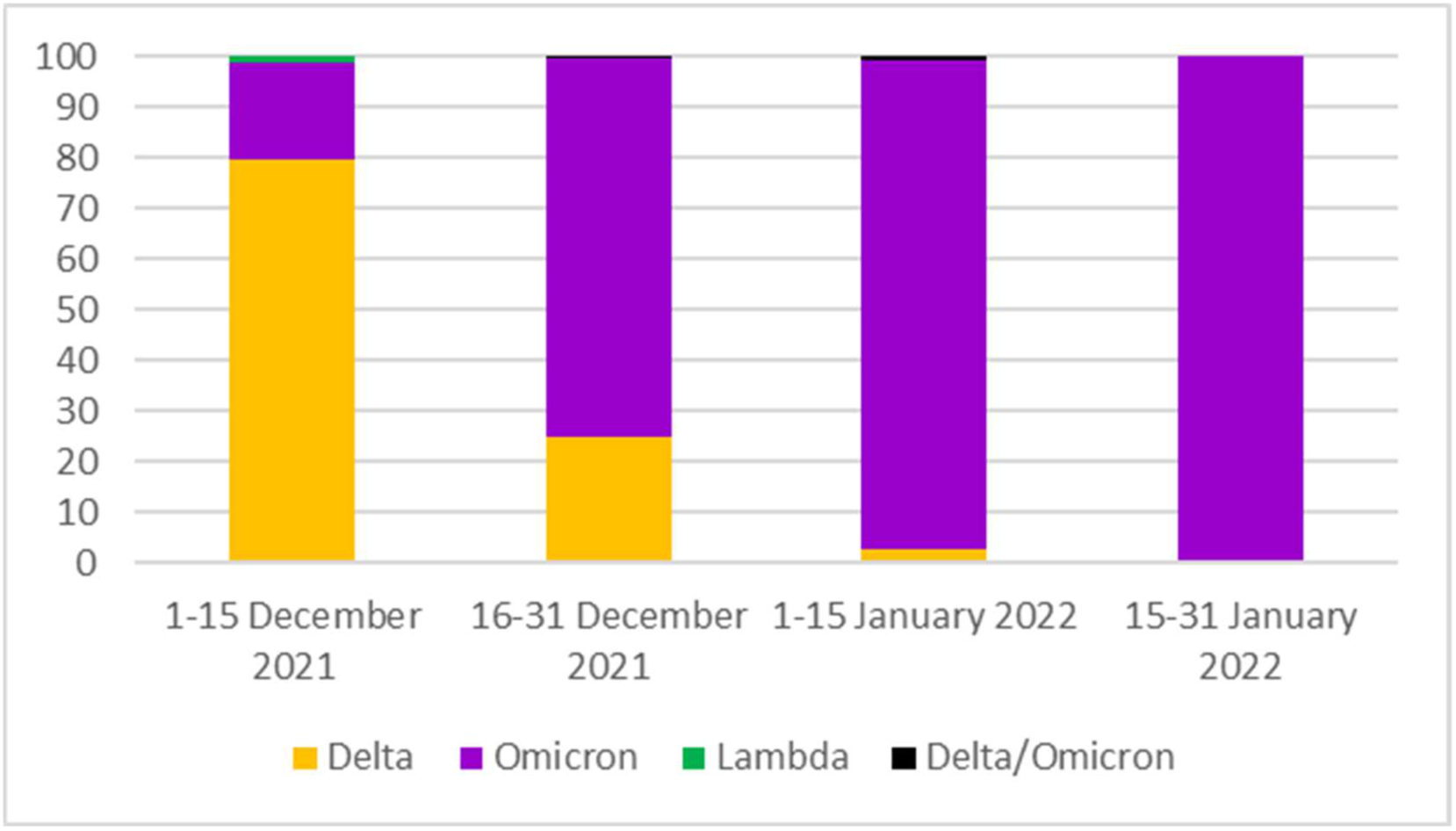
Distribution of circulating VOC/VOIs during December 2021 and January 2022 in the province of Cordoba, Argentina.

Main features of the 3 co-infected patients are shown in Table 1. Figure 2A shows profiles obtained for relevant mutations tested by real time RT-PCRs of these samples, compatible with co-infections. Detection of L452R mutation presented amplification for both, the mutation sequence as well as the wt sequence (Figure 2A, Supplementary Table 1).

**Table 1.** Main characteristics of the patients with SARS-CoV-2 Delta/Omicron co-infections.

| Sample ID | Date of swab collection/onset of symptoms | Sex | Age range (years) | Vaccine | Co-morbidities | Clinic manifestations | Hospitalization |
| --- | --- | --- | --- | --- | --- | --- | --- |
| 1 | December 2021 | M | 16-20 | Yes (complete scheme) | No | Fever, throat pain. | No |
| 2 | January 2022 | F | 0-5 | Yes (one dose) | No | Fever, dyspnea | Yes |
| 3 | December 2021 | M | 60-65 | No | Arterial hypertension | Asthenia, dyspnea, pneumonia | Yes |

**Figure 2.**
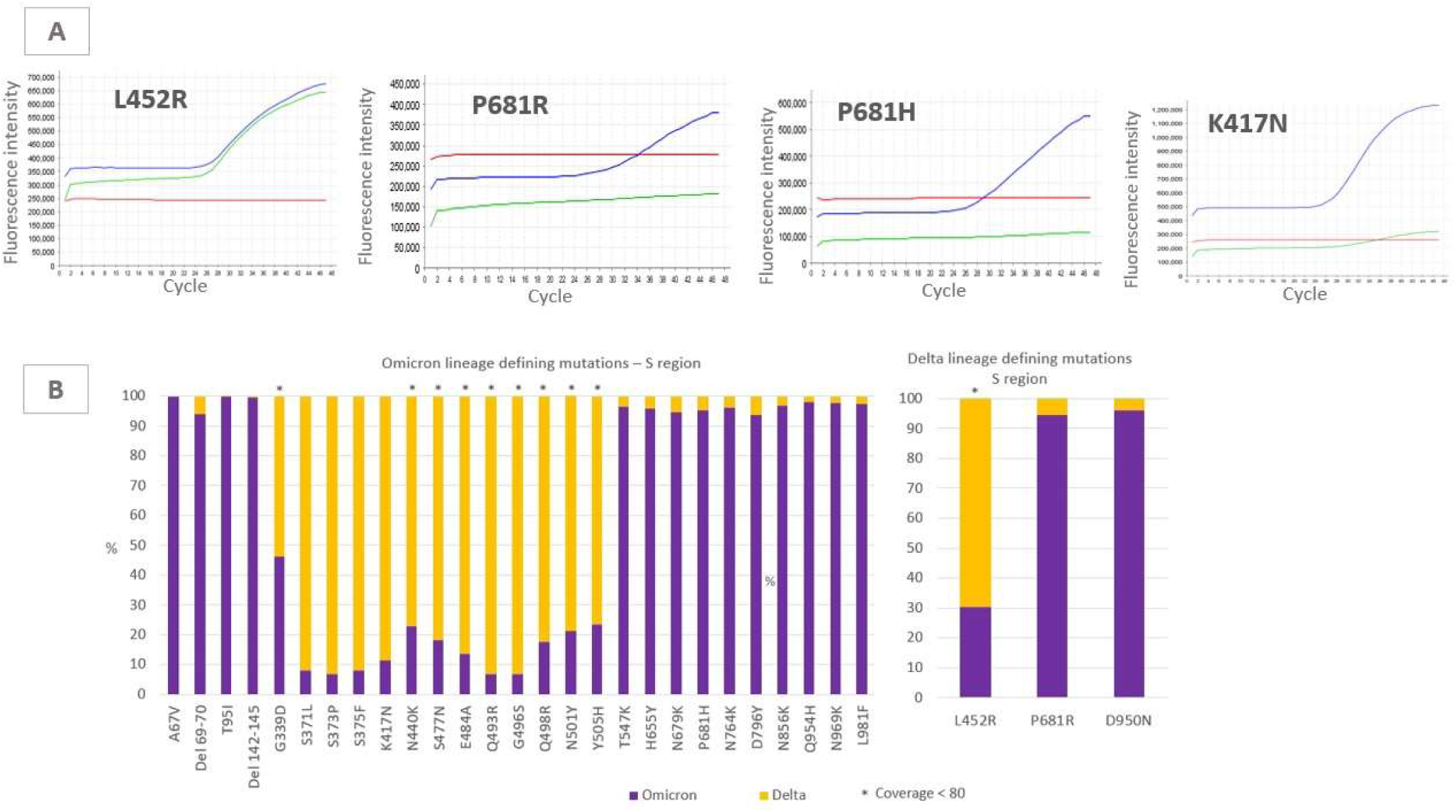
A-Real-time RT-PCR for VOC typing. Blue curve: presence of mutation; green curve: absence of mutation (presence of wild type); red curve: negative control. B)- Proportion of sequencing reads obtained for the S region by the whole genome sequencing technique matching with VOCs Omicron or Delta for the analyzed mutations that define the lineage. *Positions in which the number of reads were less than 80.

Amplification of P681R resulted positive for the mutation, but negative for the wt sequence (Figure 2A, Supplementary Table 1), which could indicate the presence of another mutation in that position. Mutation of P681H resulted positive for the mutation and negative for wt (Figure 2A, Supplementary Table 1). Finally, the mutation K417N was detected, together with the wt sequence, which was weakly amplified in the three cases (Figure 2A, Supplementary Table 1). To rule out possible cross-contaminations, the 3 samples were repeated from the original samples (nucleic acid extraction and specific RT-PCRs for VOC/VOI detection was performed), arriving at the same result.

Sample N°1 presented sufficient amount of RNA and high viral load, so the whole genome sequence could be generated for further investigation. The sequence obtained contained 29867 nucleotides, from which 0.87% were Ns and 0.34% were mutations (compared to the reference sequence WIV04). The average percentage of reads matching VOC Omicron was higher than reads matching VOC Delta (Figure 2B). Positions in which Delta was majority presented low coverage. Pangolin COVID-19 Lineage Assigner (Pangolin v3.1.19) could not assign a lineage to this sequence.

## Discussion

Coinfection with distinct SARS-CoV-2 lineages is considered a rare phenomenon. However, it is likely and is thought to be underestimated (Dezordi et al. 2021).

In this work we report the coinfection with Delta and Omicron VOCs. This is the first description of co-infected individuals carrying two distinct lineages of SARS-CoV-2 in Argentina. VOC Delta was first described in our province on July 2021, when it was detected in a traveler and his close contacts. Due to the efforts carried out by the health authorities of the province, which included tracking and isolating Delta positive cases and its close contacts, the spread of this VOC was delayed, so its increase was gradual, until reaching its highest proportion of circulation (85%) in November 2021 (Castro et al. 2021), but without a substantial increase in the number of cases (Gobierno de la provincia de Córdoba 2022).

VOC Omicron was detected in Argentina -and particularly in Córdoba province-the first days of December 2021, in a traveler from Dubai, and it quickly spread throughout the province (Gobierno de la provincia de Córdoba 2022). The sharp increase in Omicron frequency was accompanied by an increase in the number of cases, giving rise to the third wave of COVID-19 in the province and the entire country (Gobierno de la provincia de Córdoba 2022, Ministerio de Salud de la Nacion 2022).

In this context of co-circulation of variants, 3 samples with Delta/Omicron SARS-CoV-2 co-infection were identified, all of them detected the last 2 weeks of December, when co-circulation of Delta and Omicron was registered (Gobierno de la provincia de Córdoba 2022). Co-infection events between dominant SARS-CoV-2 lineages have been previously reported, also in a very low proportion of the tested samples (Dezordi et al. 2021, Hosch et al. 2022, Zhou et al. 2021). Although these events are rare, they are believed to be quite common during periods of high viral prevalence (Jackson et al. 2021) and are believed to be underreported (Dezordi et al. 2021), as they are not easy to identify. Generally, one of the lineages is present in a greater proportion (Hosch et al. 2022), which sometimes causes only one lineage to be detected. In addition, specialized personnel are required for the interpretation of the variant-specific real-time PCR technique, which is not always capable of detecting subtleties in the reaction results. In our study, co-infections were detected during the third wave of COVID-19 that took place in our country, with very high levels of viral circulation, in accordance with previous reports (Jackson et al. 2021, Dezordi et al. 2021).

The report of co-infections of SARS-CoV-2, both locally and globally, becomes relevant in a context of changing circulation of variants and emergence of new ones. In this sense, recombination, already reported for other Coronaviruses and also recently for SARS-CoV-2, is a possibility in individuals simultaneously infected with more than one lineage (Hosch et al. 2022, Jackson et al. 2021). In turn, the emergence of newly recombined viruses might result in increased transmissibility or immune evasion (Hosch et al. 2022), as recombination permits the combination of advantageous mutations from distinct variants (Jackson et al. 2021). Since recombination is only possible with co-infection, decreasing the prevalence and circulation of SARS-CoV-2 will minimize the chance of forming recombinant lineages with genetic combinations that could potentially increase virus fitness (Jackson et al. 2021).

Until now, no major clinical implications have been described in patients with co-infection with more than one SARS-CoV-2 lineage (Hosch et al. 2022). In this study, only one of the patients presented severe symptoms (pneumonia, dyspnea), although they were probably due to the fact that he was not vaccinated and to the presence of risk factors (over 60 years of age, arterial hypertension) rather than the co-infection. However, more clinical research is needed and should be carried out on these patients.

In conclusion, we found, for the first time in Argentina, co-infections by two SARS-CoV-2 lineages (Delta/Omicron) during the third wave of COVID-19, the largest in our country (Ministerio de Salud de la Nacion 2022). This highlights the importance of continuing molecular surveillance, especially in moments of high viral circulation, to detect both co-infections as well as recombinations. It is also important to continue studying clinical cases related to coinfections, in order to elucidate possible severe cases.

## Data Availability

All data produced in the present work are contained in the manuscript.

## Acknowledgements

All authors have seen and approved the manuscript.

## Ethics

The Government of the Province of Córdoba through the Ministry of Health determined that the ethical review, approval and written informed consent are not required for the study of oropharyngeal swab samples obtained from human participants in the study “SARS-CoV-2 genomic surveillance enables the identification of Delta/Omicron co-infections in Argentina” in accordance with local legislation and institutional requirements. Furthermore, the data of this study were openly available to the public before the initiation of the study.

## Competing interests

None.

## Notes

### Competing Interest Statement

The authors have declared no competing interest.

### Funding Statement

This study was funded by Ministerio de Salud de la Provincia de Cordoba, Ministerio de Salud de la Nacion Argentina and PAIS consortium.

### Author Declarations

The Government of the Province of Cordoba through the Ministry of Health determined that the ethical review, approval and written informed consent are not required for the study of oropharyngeal swab samples obtained from human participants in the study SARS-CoV-2 genomic surveillance enables the identification of Delta-Omicron co-infections in Argentina in accordance with local legislation and institutional requirements. Furthermore, the data of this study were openly available to the public before the initiation of the study.

