## Supplemental Table 1 for "SARS-CoV-2 genomic surveillance enables the identification of Delta/Omicron co-infections in Argentina"

**Supplementary Table 1**. Results and Ct values of the mutation-specific real time RT-PCRs tested.

| **Sample ID** | **Ct GENE N (SARS-CoV-2 amplification)** | **L452R** | | **K417N** | | **P681R** | | **P681H** | |
| --- | --- | --- | --- | --- | --- | --- | --- | --- | --- |
|  |  | **WT** | **Mut** | **WT** | **Mut** | **WT** | **Mut** | **WT** | **Mut** |
| 1 | 20.6 | 23.9 | 26.9 | 26.6 | 26 | Negative | 30 | Negative | 23.7 |
| 2 | 28.4 | 31.3 | 34 | 32.2 | 31.6 | Negative | 39 | Negative | ND* |
| 3 | 27.5 | 33.4 | 34.8 | 36.6 | 36.8 | Negative | 41 | Negative | 32 |

*No data. Ct: threshold cycle. WT: wildtype. Mut: mutation.
